# Postprandial dynamics of proglucagon cleavage products and their relation to metabolic health

**DOI:** 10.1101/2021.11.25.21266733

**Authors:** Robert Wagner, Sabine S. Eckstein, Louise Fritsche, Katsiaryna Prystupa, Sebastian Hörber, Hans-Ulrich Häring, Andreas L. Birkenfeld, Andreas Peter, Andreas Fritsche, Martin Heni

**Affiliations:** Institute for Diabetes Research and Metabolic Diseases of the Helmholtz Center Munich at the University of Tübingen, Tübingen, Germany; German Center for Diabetes Research (DZD), Tübingen, Germany; Department of Internal Medicine IV, Division of Diabetology, Endocrinology, and Nephrology, Eberhard Karls University Tübingen, Tübingen, Germany; Institute for Clinical Chemistry and Pathobiochemistry, Department for Diagnostic Laboratory Medicine, University Hospital Tübingen, Tübingen, Germany

## Abstract

While oral glucose ingestion typically leads to a decrease in circulating glucagon levels, a substantial number of persons display stable or rising glucagon concentrations when assessed by radioimmunoassay (RIA). However, these assays show cross-reactivity to other proglucagon cleavage products. Recently, more specific assays became available, therefore we systematically assessed glucagon and other proglucagon cleavage products and their relation to metabolic health. We used samples from 52 oral glucose tolerance tests (OGTT) that were randomly selected from an extensively phenotyped study cohort.

Glucagon concentrations quantified with RIA were non-suppressed at 2 hours of the OGTT in 36 % of the samples. *Non-suppressors* showed lower fasting glucagon levels compared to *suppressors* (p=0.011). Similar to RIA measurements, ELISA-derived fasting glucagon was lower in *non*-*suppressors* (p<0.001). Glucagon 1-61 as well as glicentin kinetics were significantly different between *suppressors* and *non*-*suppressors* (p=0.004 and p=0.002, respectively) with higher concentrations of both hormones in non-*suppressors*. Levels of insulin, C-peptide, and free fatty acids were comparable between groups. Non*-suppressors* were leaner and had lower plasma glucose concentrations (p=0.03 and p=0.047, respectively). Despite comparable liver fat content and insulin sensitivity (p≥0.3), they had lower 2-hour post-challenge glucose (p=0.01).

Glucagon 1-61, glicentin and GLP-1 partially account for RIA-derived glucagon measurements due to cross-reactivity of the assay. However, this contribution is small, since the investigated proglucagon cleavage products contribute less than 10% to the variation in RIA measured glucagon. Altered glucagon levels and higher post-challenge incretins are associated with a healthier metabolic phenotype that is known to be indicative for reduced cardiovascular risk, cancer incidence, and mortality.

## Introduction

Glucagon, glicentin, and glucagon-like peptide (GLP)-1 as proglucagon cleavage products all originate from the same preproglucagon gene (see figure 1). Differential expression of the preproglucagon gene in varying tissues is accompanied by differential processing of the proglucagon transcript by prohormone convertases.

**Figure 1.**
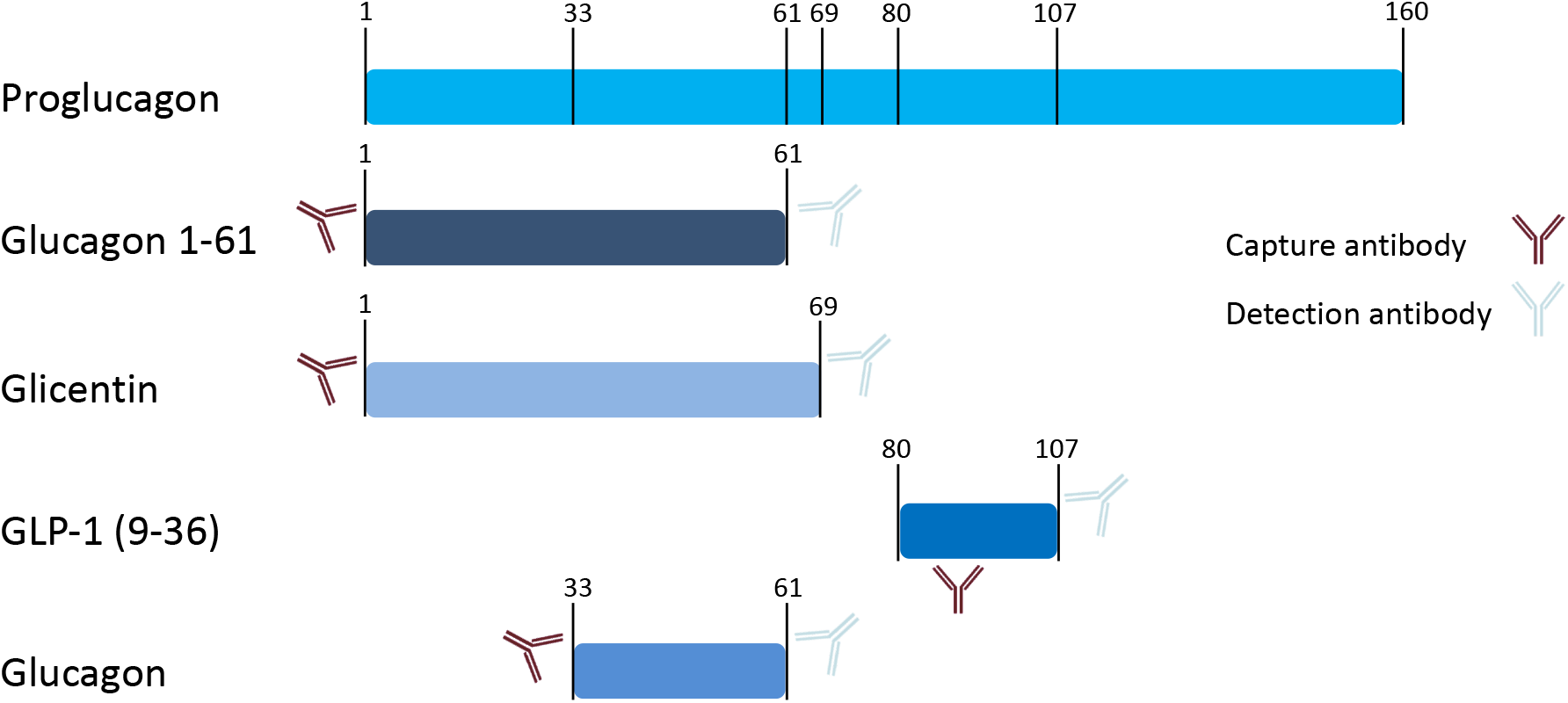
Schematic presentation of proglucagon and proglucagon cleavage products. Numbers indicate amino acid positions of cleavage sites. Antibodies schematically indicates epitopes that are used by the commercial immunoassays applied in our measurements (as provided by the manufacturer).

For a long time, counteracting insulin was thought to be the main function of glucagon. Since patients with type 2 diabetes show elevated glucagon levels^1^, it has been believed to be a relevant contributor to hyperglycemia. Research over the last years, however, revealed a much more complex role of glucagon with potentially beneficial effects for whole-body metabolism^2,3^.

GLP-1 is an insulinotropic peptide that potentiates insulin secretion and mainly originates from the L cells in the distal small bowel and colon^4,5^. Glicentin is also produced in the intestinal L cells and its exact functions are still enigmatic. Studies in animal models and *in vitro* human tissues suggest a regulatory function on intestine, gastric acid production and insulin production^6–12^.

Studies have shown that bariatric surgery for the treatment of obesity results in dramatic metabolic changes which can at least partially be attributed to incretins such as GLP-1 and glicentin^13–15^. Uncovering the complex regulation that underlies the beneficial effects of bariatric surgery requires a better understanding of different incretins and their interplay on metabolism.

In a multi-cohort study with more than 4000 participants who underwent oral glucose tolerance tests, we found non-suppressed glucagon levels at 2-hours after glucose load in 21-34% of the study populations^16^. Surprisingly, this non-suppression of glucagon was associated with a metabolically healthier phenotype. One limitation of the findings was the radioimmunoassay used to quantify glucagon in the study as this assay is known for cross-reactivity with other proglucagon cleavage products^17^. This is caused by the overlapping amino acid sequences of glucagon with other incretin peptides, resulting in assay cross-reactivity when the capture antibody binds to an epitope within these shared sequences. The proglucagon sequence from amino acid position 33-61 corresponding to glucagon. The alternative gene product glicentin spans amino acids 1-69 of the proglucagon gene, whereas GLP-1 comprises amino acids 80-107 (figure 1). Albrechtsen et al. investigated glucagon 1-61, which is another circulating proglucagon fragment. Glucagon 1-61 has been shown to stimulate insulin secretion and act on human hepatocytes in a series of experiments ^18^. Glucagon 1 – 61 comprises the amino acid sequence of glucagon and glicentin but lacks the eight C-terminal amino acids of glicentin^18^ (figure 1).

Due to a cross-reactivity of the radioimmunoassay, it is plausible that the positive effects of glucagon on metabolism we observed in our previous study result from effects of other proglucagon fragments. Therefore, we aimed to investigate the relationship of glucagon before and after an OGTT with other proglucagon fragments using a highly specific glucagon immunoassay.

## Results

Glucagon concentrations were first measured by radioimmunoassay (figure 2A). Analyzing these measurements, 36 % of the participants showed stable or rising RIA-derived glucagon concentrations during the OGTT. We defined them as *non*-*suppressors*. Of note, the *non-suppressors* had significantly lower fasting levels of glucagon compared to the *suppressors* (p=0.011). In parallel, samples were measured with the highly specific glucagon ELISA, showing lower values compared to RIA. Bland-Altman plots revealed reasonable agreement between the two methods with on average 15±6 pmol/l higher concentrations from RIA measurements (figure 3).

**Figure 2.**
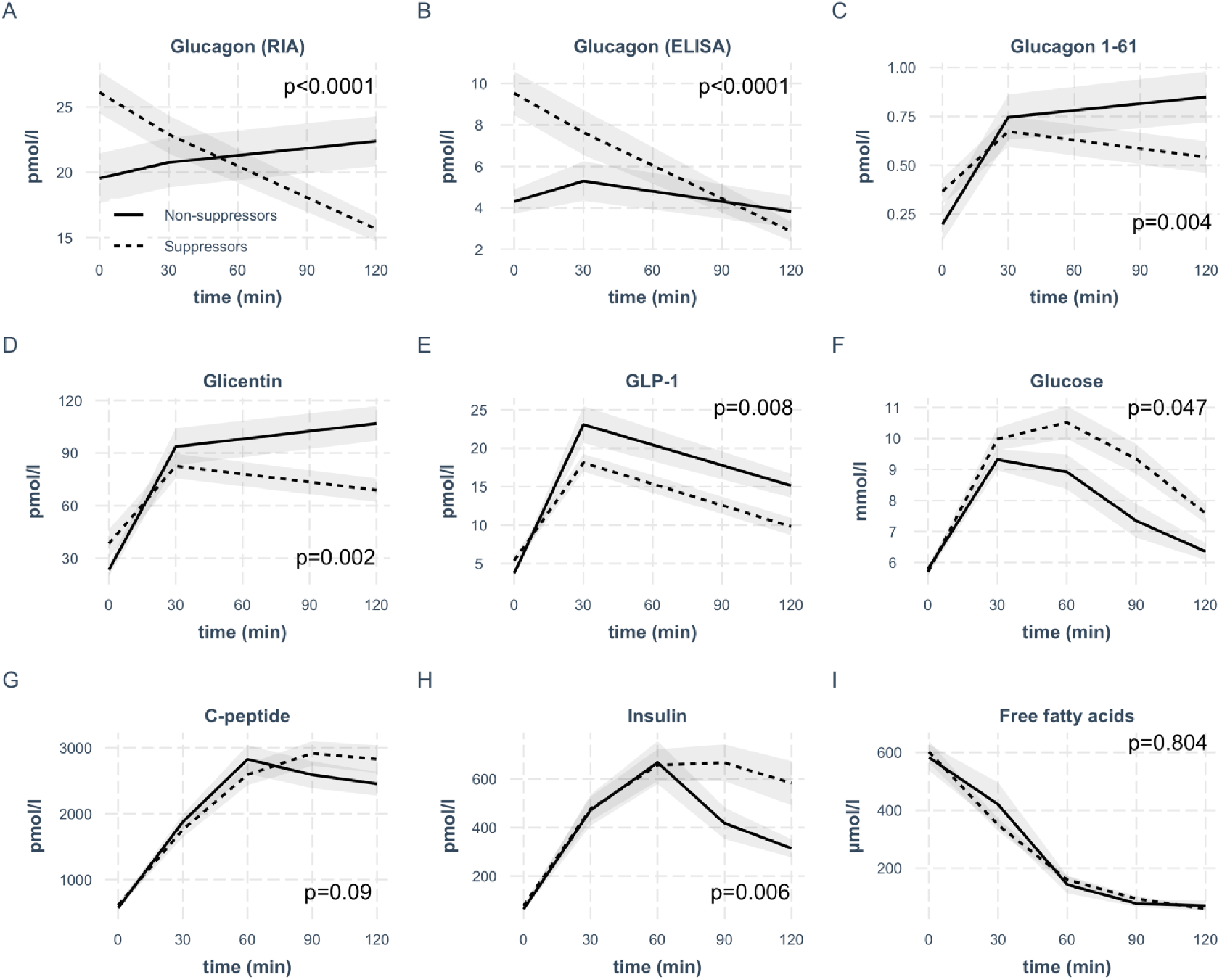
Concentrations of investigated analytes in the groups of glucagon *suppressors* and *non*-*suppressors* during the OGTT. The respective analyte is indicated in the box. Lines represent means with standard errors, p-values are calculated using linear mixed model. N=52.

**Figure 3.**
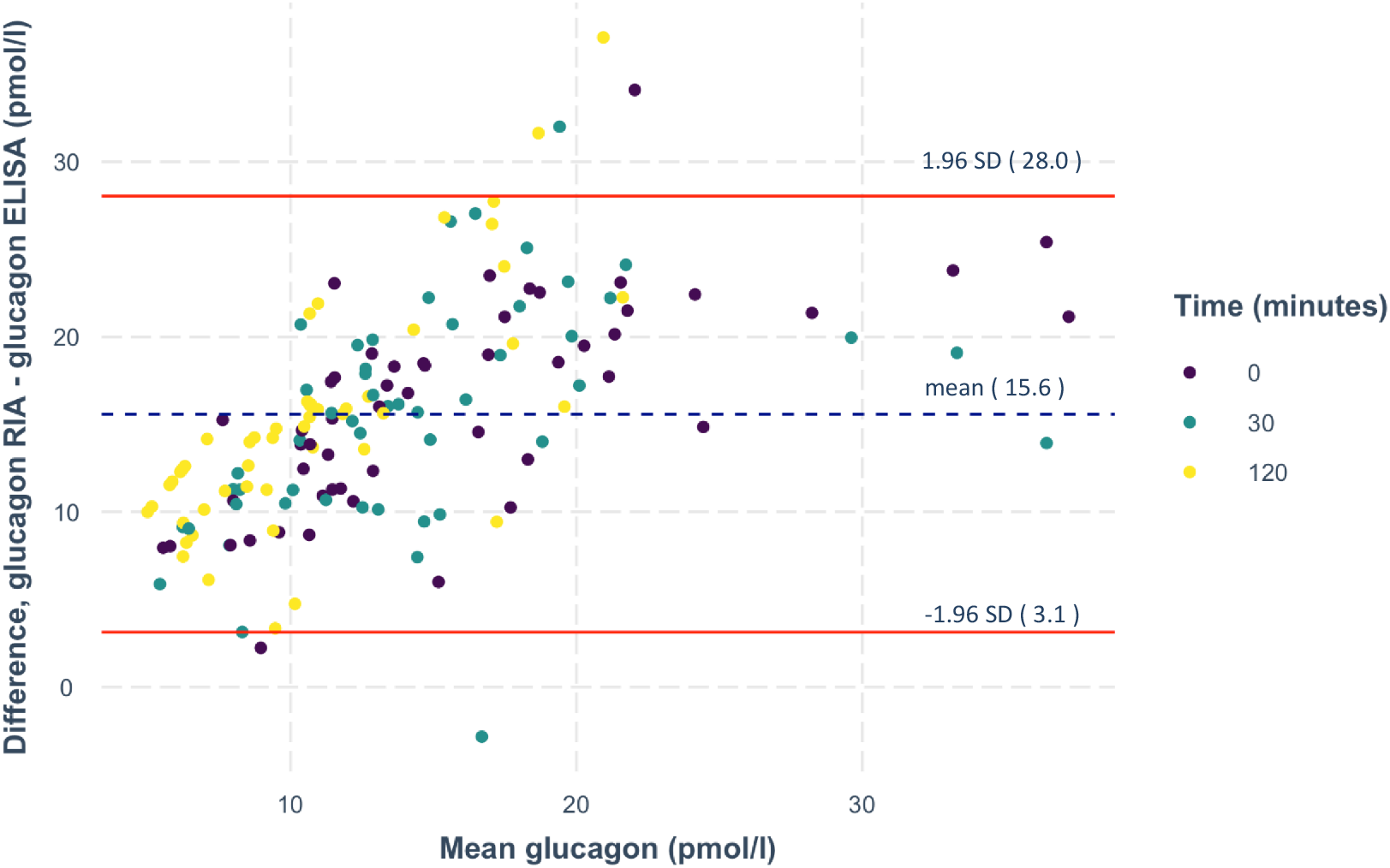
Bland-Altman plot of the RIA- and ELISA-measured glucagon. Differences in glucagon measurements between the two assays are plotted against mean of glucagon values. The dashed lines represent the mean, the solid lines depict the lines of agreement calculated as mean ± 1.96 times the SD of this difference. N=156 measurements from 52 oGTTs.

The ELISA also detected persons with stable or rising glucagon. Similar to RIA measurements, fasting glucagon was lower in the *non*-*suppressors* (p<0.001). The RIA- and ELISA-derived glucagon *suppressors* and *non*-*suppressors* showed similar kinetics, regardless of which assay the hormone was measured with (figure 2 A and B). As the applied glucagon RIA is known to have cross-reactivity with other proglucagon cleavage products^19,20^, we quantified these hormones by highly specific ELISAs. We measured glucagon 1-61, glicentin and GLP-1 from the same samples (figure 2 C-E).

To investigate the relative contribution of these proglucagon cleavage products to RIA-derived glucagon measurements, we modeled glucagon RIA levels at all available OGTT time points using glucagon ELISA, glicentin, glucagon 1-61 and GLP-1. The share of total variance explained by this model was 82.2%. The combination of the above-mentioned variables explained 43.6% as fixed effects. By removing each factor separately, we determined their relative contributions. In figure 4, we show the relative contribution of glicentin, GLP-1 and glucagon to the total variance (total variance was set to 100%). ELISA measured glucagon explained 93% of the variance, whereas glicentin explained 5% of it. The contribution of GLP-1 (2%) was not statistically significant. Inclusion of Glucagon1-61 in the model deteriorated its performance and has been therefore removed from the final model.

**Figure 4.**
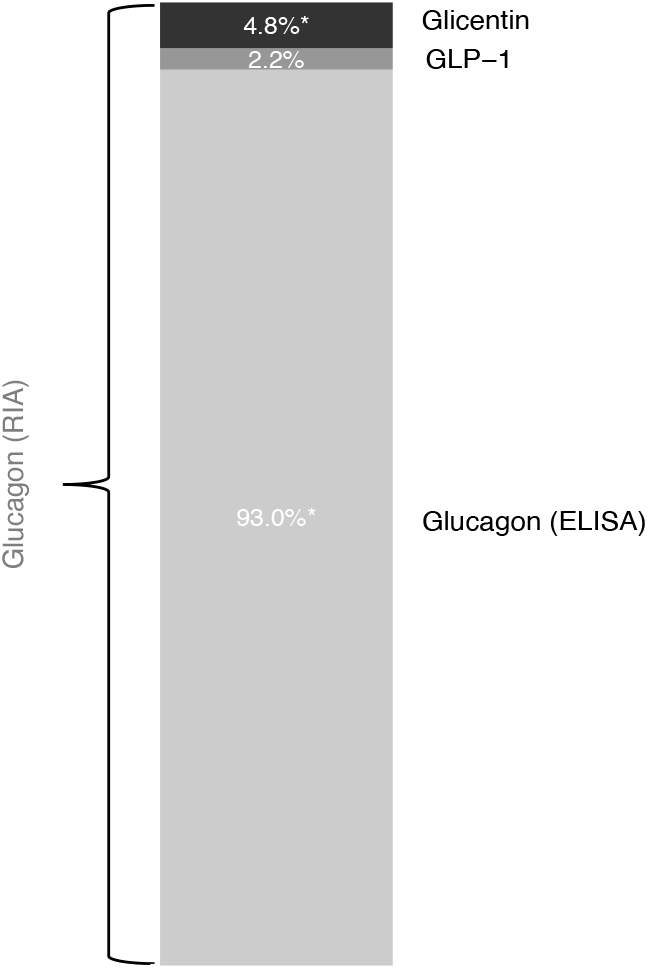
Relative contribution of proglucagon cleavage products to RIA-derived glucagon measurements. We used linear mixed models with the participant as random intercept and the OGTT-time point as fixed effects. Marginal R squared was calculated to describe the proportion of variance in the outcome variable explained by the fixed effect. Removing each factor separately, we determined the percentage of its respective contribution to RIA-derived glucagon measurements that are presented here as a bar chart.

While the fasting concentration of glucagon 1-61 and glicentin were comparable between groups (p≥0.1), GLP-1 was significantly lower in *non*-*suppressors* (p=0.02).

Glucagon 1-61 as well as glicentin kinetics were significantly different between *suppressors* and *non*-*suppressors* with higher concentrations of both hormones in *non-suppressors* (figure 2C/D). For GLP-1, the concentrations were not significantly different (figure 2E).

As incretins stimulate pancreatic insulin release, we next analyzed insulin and C-peptide levels as well as plasma glucose. Insulin and C-peptide levels were comparable between the two groups (figure 2 G/H). However, insulin concentrations were lower in the *non*-*suppressors* in the last 30 minutes of the OGTT, resulting in a statistical difference between groups. In line with comparable insulin secretion, suppression of free fatty acids was not different between groups (figure 2I). However, plasma glucose concentrations were significantly lower in the *non*-*suppressors* (figure 2F), even after adjustment for BMI (p=0.04).

The *non*-*suppressors* were leaner (p=0.03) and had a lower waist circumference (p=0.04). Despite comparable liver fat content and insulin sensitivity (p≥0.3), they had lower post-challenge glucose concentrations (p=0.01), which remained significant after adjustment for BMI (p=0.002).

## Discussion

In a meta-analysis with over 4000 participants, we previously identified non-suppression of glucagon in response to oral glucose intake, to be linked to a favorable whole-body metabolism^16^. One limitation of that study was quantification of glucagon by RIA that likely cross-reacts with additional proglucagon cleavage products. We therefore performed comparative analyses of glucagon kinetics assessed by RIA and a highly sensitive ELISA. In addition, we investigated the kinetics of the major proglucagon cleavage products glucagon 1-61, glicentin and GLP-1. Regardless of the measurement approach, we identified persons with glucagon suppression during the OGTT and such with stable or even rising concentrations. The persons that did not suppress RIA-measured glucagon in response to oral glucose intake showed higher incretin responses during the glucose challenge. Of note, these differences in glucagon and incretin kinetics were accompanied by a metabolically healthier phenotype.

Our data suggest that glucagon 1-61, glicentin and GLP-1 partially account for RIA-derived glucagon measurements. However, this contribution is small, because *(i)* there is a reasonable agreement between RIA and ELISA measurements and *(ii)* the investigated proglucagon cleavage products explain less than 10% of total glucagon measured with RIA.

There is substantial controversy on the role of glucagon in the pathogenesis of diabetes^21,22^. While glucagon promotes hepatic glucose production, this effect is self-limited, and compensated by glucagon’s stimulation of insulin secretion, inhibition of hepatic triglyceride synthesis, increase in basal energy expenditure and central inhibitory effects on appetite^3,22,23^. Glucagon also stimulates GLP-1 receptors, though, to a lesser extent than its primary ligand^24^, that has been proven beneficial in the treatment of both diabetes and obesity. Thus, although there is an association of fasting hyperglucagonemia and type 2 diabetes^25,26^, this might be secondary to compensatory processes without playing a causal role in diabetes development. When it comes to postprandial changes in glucagon concentrations, results are even more puzzling^16,27–29^ and the detailed contribution in the pathogenesis of type 2 diabetes is still unclear^21^.

In our current study, higher post-load levels of glucagon 1-61, glicentin and GLP-1 in the non-suppressor group are associated with lower blood glucose levels. Given the insulinotropic effect of incretins, the most obvious explanation could be a difference in insulin secretion between the two groups. However, our data argue against this, as we detected no significant difference in C-peptide levels, which are indicative for pancreatic insulin secretion. This is further underlined by comparable insulin-induced suppression of lipolysis, which was assessed by free fatty acid concentrations.

Fasting glucose, that strongly relies on endogenous glucose production^30^ is not different between *suppressors* and *non*-*suppressors* glucagon groups. This suggests that endogenous glucose production does not have a relevant contribution to differences in glycaemia between the two groups. Comparable whole-body insulin sensitivity argues against major differences in hepatic insulin sensitivity that would result in different suppression of endogenous glucose production during the OGTT.

One possible explanation for the altered glucose levels is therefore a difference in non-insulin dependent glucose disposal. Non-insulin dependent glucose disposal is also referred to as glucose sensitivity or glucose effectiveness. This term describes the ability of glucose to regulate glucose disposal and gluconeogenesis by itself under basal insulin conditions^31^. Tissues such as fat and muscle usually take up glucose in an insulin-dependent fashion. However, insulin-independent glucose uptake is also present^32,33^. Glucagon has direct glucoregulatory effects through the brain^23^. Thus, one possibility is that glucose effectiveness is regulated via the brain, though the molecular mechanisms are still unclear^34^. Our results suggest that glucagon or incretins could be involved and may promote non-insulin dependent glucose disposal. This mechanism could contribute to the glucose-lowering effect of these hormones and to recent pharmacotherapies that target these pathways.

Our study is limited by the sample size and the fact that we did not apply mass spectrometry as gold standard for the identification of peptide hormones. In addition, we did not test additional stimuli that potentially trigger gastrointestinal secretion of proglucagon cleavage products, e.g. mixed meal test.

In summary, we verified that in some persons oral glucose intake does not result in a suppression of glucagon. Of note, these persons additionally displayed higher post-load concentrations of further preproglucagon cleavage-products, including glicentin. As these persons are leaner and have lower blood glucose, our results indicate that proglucagon cleavage-products could contribute to the development of a healthier metabolic phenotype that is known to be indicative for reduced cardiovascular risk, cancer incidence, and mortality.

## Methods

### Subject characteristics and measurements

Samples from 52 randomly selected participants of the prediabetes lifestyle intervention study^35^ (PLIS, registered with clinicaltrial.gov NCT01947595) were investigated in this project. None of the study participants took any kind of medication that interferes with glucose or energy metabolism. The study was approved by the local ethics board (Ethics Committee of the Medical Faculty of the Eberhard Karls University and the University Hospital Tübingen) and conducted in accordance with the declaration of Helsinki. All participants provided written informed consent.

Oral glucose tolerance tests (OGTT) were performed after an overnight fasting period of 12 hours. Venous blood was drawn at baseline and 30, 60, 90 and 120 minutes after drinking a 75 g glucose solution (Accu-Check Dextrose O.G.T., Roche Diagnostics, Mannheim, Germany). Plasma glucose and free fatty acids were measured from sodium fluoride plasma with an ADVIA chemistry XPT autoanalyzer (Siemens Healthineers). Serum insulin and C-peptide were analyzed using ADVIA Centaur XPT immunoassay system (Siemens Healthineers). Study participants were categorized into glycemic groups: normal fasting glucose, impaired fasting glucose, impaired glucose tolerance, impaired fasting glucose + impaired glucose tolerance, and type 2 diabetes according to ADA criteria^36^. For details see table 1.

**Table 1.** Subject characteristics.

|  | Stratified by glucagon RIA dynamics |  |  |  |
| --- | --- | --- | --- | --- |
|  | Suppressors | Non-suppressors | P<sub>unadjusted</sub> | <sup>1</sup>P<sub>adjusted</sub> |
| n | 33 | 19 |  |  |
| Age (years) | 59 (9) | 61 (11) | 0.397 |  |
| BMI (kg/m<sup>2</sup>) | 29.3 (5.1) | 26.5 (3.4) | 0.034* |  |
| Sex (f/m) | 14/19 | 9/10 | 0.956 |  |
| <sup>2</sup>NFG/IFG/IGT/IFG+IGT/DIA | 9/10/3/7/4 | 6/10/0/2/1 | 0.365 |  |
| Insulin sensitivity index, OGTT-derived (AU) | 10.9 (9.7) | 12.0 (5.6) | 0.633 | 0.572 |
| <sup>3</sup>Liver fat content (%) | 6.8 (8.0) | 4.9 (4.7) | 0.389 | 0.544 |
| HbA1c (%) | 5.8 (0.5) | 5.7 (0.3) | 0.499 | 0.832 |
| Fasting glucose (mmol/l) | 5.7 (0.63) | 5.8 (0.76) | 0.635 | 0.376 |
| 2-hour glucose (mmol/l) | 7.6 (1.8) | 6.4 (1.2) | 0.010* | 0.002* |
| Waist circumference (cm) | 100.0 (15.9) | 91.3 (9.7) | 0.035* | 0.566 |
| Hip circumference (cm) | 105.9 (9.8) | 101.3 (7.2) | 0.077 | 0.981 |
| Waist-to-Hip-Ratio (WHR) | 0.94 (0.1) | 0.90 (0.1) | 0.129 | 0.613 |

Numbers in columns represent mean and standard deviation
<sup>1</sup>adjusted for BMI and sex
<sup>2</sup>Normal fasting glucose (NFG), impaired fasting glucose (IFG), impaired glucose tolerance (IGT), participants with T2D (DIA)
<sup>3</sup>Total measurements are 44, for "Suppressors" n=28, for "Non-suppressor" n=16

Glucagon was measured by a commercially available radioimmunoassay (Linco Research/Millipore, St. Charles, MO). Glucagon, glicentin and GLP-1 were also measured with commercially available ELISA assays (Mercodia, Uppsala, Sweden) according to the manufacturer’s instructions. The antibody binding sites^18,37^ are indicated in figure 1. Glucagon 1-61 was measured with a prototype ELISA from Mercodia (Uppsala, Sweden) according to the manufacturer’s instructions. The prototype ELISA shows 0.5 % cross-reactivity with glicentin and < 0.2 % with glucagon respectively. To ensure optimal sample quality, EDTA plasma was stabilized with 300 ng/ml protease inhibitor aprotinin (Sigma, Merck, Darmstadt, Germany) und subsequently processed at 4°C and kept frozen at -80°C until batch measurement.

HbA1c was measured by HPLC. Height, weight, waist and hip circumference were measured according to standard operating procedures. Liver fat content was measured by localized ^1^H-MR spectroscopy using a 1.5 T MR scanner (Magnetom Sonata, Siemens Healthcare, Erlangen, Germany) and the distribution of lean and adipose tissues was measured by whole body MR imaging as previously described^38^.

### Calculations

Insulin sensitivity was assessed with the index proposed by Matsuda and De Fronzo^39^.

### Statistics

The relative change of glucagon RIA from 0 to 120 min was used to define the groups of *suppressors* and *non*-*suppressors*. The *suppressors* were determined as participants with the fold change glucagon RIA (glucagon RIA 120 min/glucagon RIA 0 min) less than 1, while *non*-*suppressors* were those with equal or higher than 1.

To analyze measurements from the same participants at different time-points of the OGTT, linear mixed models with the participant as random intercept and the OGTT-time point as fixed effects were used. Marginal R squared was calculated to describe the proportion of variance in the outcome variable explained by the fixed effect. Removing each factor separately, we determined the percentage of its respective contribution to RIA-derived glucagon measurements.

The Wilcoxon rank-sum/Kruskal-Wallis test was used to perform nonparametric tests on continuous variables, for comparing two independent samples. Categorical variables were compared by chi-squared test. P-values < 0.05 were considered statistically significant. Statistical analyses were performed with R (software version 4.0.3)^40^.

## Data Availability

All data produced in the present study are available upon reasonable request to the authors

## Acknowledgments

We gratefully acknowledge the excellent technical assistance of Dorothee Neuscheler and Henrike Peuker (both University Hospital Tübingen). We thank Dr. Monica Vilhelmsson (Mercodia, Uppsala, Sweden) for scientific discussions about specificity and sensitivity and about the antibody binding areas of the used ELISAs. We furthermore thank Mercodia (Uppsala, Sweden) for making their prototype glucagon 1-61 ELISA available for purchase and for technical input.

## Author contributions

RW, SES, LF and MH designed the analysis strategy, supervised the project and contributed to discussion. SES, LF, SH and AP performed measurements and contributed to discussion. LF and KP did statistical analysis. HUH, ALB and AF contributed to discussion. RW, SES and MH drafted the manuscript. All authors approved the final version of the manuscript prior to submission.

